# Designing a Structural Equation Model (SEM) for the Relationship between Quality of Working Life and Job Involvement among Nurses, with the Mediating Role of Job Burnout and Job Identity

**DOI:** 10.1101/2024.11.12.24317140

**Authors:** Elham Nazari, Azin Roumi, Toomaj Sabooteh

## Abstract

**Introduction:** Given the issue of nursing shortages globally and in Iran, compounded by new global conditions due to the COVID-19 pandemic, the quality of working life (QWL) among nurses and the factors influencing it have become important issues. Therefore, this study aimed to investigate the relationship between QWL and job involvement among nurses, considering the mediating role of job burnout and job identity.

**Methodology:** This study is a cross-sectional analysis conducted in Khorramabad, Lorestan province, Iran. The statistical population comprised nurses employed in Khorramabad hospitals. The sample size was determined using Cochran’s formula, and simple random sampling was employed. To assess the variables, the Work-Related Quality of Life Scale (WRQOLS), Maslach Burnout Questionnaire, Hao’s Professional Identity Questionnaire, and the Utrecht Work Engagement Scale (UWES-9) were used. Structural Equation Modeling (SEM) was employed to examine the relationships between variables, and Smart-PLS software was used for data analysis.

**Results:** Results showed a significant negative correlation between job involvement, job identity, and QWL with job burnout. The correlation coefficients were -0.910, -0.878, and -0.900, respectively. Additionally, the results indicated that job identity and job burnout mediate the relationship between QWL and job involvement among nurses.

**Conclusion:** According to the findings of this study, job identity and job burnout have a mediating effect on the relationship between QWL and employee engagement. Therefore, when nursing managers aim to enhance the job engagement of new nurses, they should consider not only QWL but also the aspects of job identity and job burnout. Enhancing QWL can improve psychological empowerment and reduce nurse burnout. Moreover, professional training and improvement of communication skills among nursing staff can assist them in managing job burnout.

## 1. Introduction

Nurses are an integral part of any healthcare system. Currently, the shortage of nurses is a global health issue. Despite significant resources and efforts dedicated to the training and recruitment of nurses, challenges within this profession persist. The effectiveness and efficiency of healthcare systems are under threat, and in the long term, this could be a significant obstacle to achieving healthcare development goals for policymakers, healthcare institutions, and patients alike (1). Therefore, improving organizational management, enhancing staff commitment to the organization, and reducing nurse turnover are vital for healthcare systems. Most countries are now facing a serious nursing shortage, and this issue is likely to persist for an extended period. Particularly in aging populations and those with a high prevalence of chronic diseases, the shortage of nurses will continue. Individuals aspiring to become nurses must complete full-time general nursing or midwifery programs for over three years at professional universities. Newly employed hospital nurses generally require two years of standardized training (2).

In recent years, standardized training for new nurses has shifted from a previous emphasis on knowledge and skills to a “demand-driven and post-nursing competency-centered core” (3). For example, some hospitals in China have initiated at least two years of standardized clinical training for new nurses. On one hand, this policy has effectively improved the professional quality and service capabilities of nurses. On the other hand, newly employed clinical nurses must not only adapt to the transition from being a student to a practicing nurse, providing high-quality nursing care, but also receive training and assessment across various departments during their free time. Furthermore, the wages for newly trained nurses are low (4). This situation presents a significant challenge for new nurses. If they cannot effectively cope with the impact of this transition, their turnover rate will range from 35% to 60% (5). In Portugal, job engagement is a critical and negative predictor of turnover intentions among nurses (6). In countries such as Italy and China, job engagement also has a significant negative impact on turnover intentions (7,8). Additionally, one study indicated that 35% to 60% of new nurses left their first place of employment within a year (9). Therefore, approaches to improving the job engagement of new nurses play a crucial role in reducing their turnover rates (2).

The quality of work life (QWL) serves as an informative indicator for providing insights into potential organizational management reforms, given its broad structure encompassing job satisfaction and other factors related to individual health and organizational goals. The key value in conceptualizing QWL is the pursuit of dual objectives: increased organizational effectiveness and improved employee quality of life (10). Consequently, assessing and monitoring the QWL of nurses can benefit healthcare organizations striving to develop strategies and initiatives aimed at retaining nurses, enhancing their job efficiency, protecting them from excessive work-related pressures (burnout), and other psychosocial workplace risks, ultimately leading to reduced human resource costs and improved patient outcomes (11).

This study examines the relationship between nurses’ QWL and job involvement, with the mediating roles of job burnout and job identity, which may contribute to the research literature in this field. The importance of this study lies in several key aspects: first, it is one of the initial studies on professional identity among nurses, thereby contributing to the research domain in the literature. Second, it develops a scale for nurses’ QWL, validated through multiple quantitative assessments with a sample of 310 employed nurses, thus adding empirical testing and scale development evidence to the focal research field. Third, this study links the construct of professional identity with workplace performance metrics, providing valuable theoretical implications for the examination of critical work criteria.

### 1.1. Job Involvement

In this study, the definition of job involvement is tailored specifically to nursing, based on the conceptual analysis conducted by Bargagliotti (12). This definition emphasizes that dynamic caregiving practices emerge from an environment of autonomy and trust, leading to safer and more cost-effective outcomes for patients. Job involvement measures positive work-related satisfaction, characterized by vigor, dedication, and absorption (13). Vigor includes a high level of energy and mental resilience at work. Dedication reflects a strong commitment to one’s work, along with a sense of meaning, enthusiasm, inspiration, pride, and challenge. Absorption is characterized by concentration, enjoyment, and a rapid passage of time, making it difficult for individuals to detach from work (13,14). Research shows that job involvement in nurses is directly linked to patient safety, quality, and a positive patient experience (14,15). Positive correlations have been found between nurses’ job involvement, patient experience, and the quality and safety of nursing care (16-19). Studies indicate that a one-unit decrease in employee engagement is associated with a 29% reduction in hospital safety levels (18).

### 1.2. Quality of Work Life and Its Impact on Job Involvement, Burnout, and Job Identity

The quality of work life (QWL) of nurses refers to the extent to which the work experience of employed nurses can meet their essential personal needs (20). QWL encompasses four main dimensions: work-family balance, workload, work environment, and social needs. Some research on employees in the telecommunications industry has shown a correlation between QWL and employees’ work commitment (21). Additionally, studies across various populations have demonstrated that the work-life quality dimension significantly correlates positively with employee job involvement (22). However, there remains a lack of empirical evidence in nursing research to illustrate the relationship between QWL and nurses’ job commitment. Research indicates that low QWL is strongly associated with job burnout (23,24). Moreover, researchers have found that QWL is a significant predictor of job burnout in nurses (25), emphasizing its critical role in predicting potential stress and burnout among employees due to limited resources and organizational support (26,27). Nonetheless, studies on the relationship between these factors remain limited (28,29).

A study on family physicians found that job identity mediates the effect of QWL on presenteeism (30). However, few studies have examined the relationship between nurses’ QWL and job identity. In numerous studies on nurses, findings have shown that job identity mediates the impact of various variables on job involvement, such as the effect of professional benefits on job involvement (31) and the impact of nurse-physician collaboration on job involvement (32). Efraty & Sirgy (1990) suggested that understanding QWL directly influences employees’ job identity (33). Thus, the first hypothesis of this study proposes that job identity mediates the relationship between QWL and job commitment:

**H1:** The QWL of nurses with standard training positively correlates with job involvement.

**H2:** QWL negatively correlates with job burnout.

**H3:** QWL positively correlates with job identity.

**H4:** Job identity mediates the relationship between QWL and job commitment.

### 1.3. Job Burnout and Its Relationship with Job involvement

The term “job burnout” was first introduced by the American clinical psychologist Freudenberger (34). In 1981, Maslach and Jackson defined job burnout as a prolonged response of individuals in the helping industry, who cannot effectively cope with persistent workplace pressures (35). This syndrome comprises emotional exhaustion, depersonalization, and a reduced sense of personal accomplishment. Job burnout is characterized by prolonged work-related stress, which negatively impacts efficiency and the quality of medical care (36). More importantly, job burnout is closely associated with higher turnover rates among nurses (37-40). Studies involving dentists, critical care physicians, and other groups have shown a significant negative correlation between job burnout and job involvement (41,42). As proposed by the Job Demand-Resources (JD-R) model, job burnout is considered one of the possible outcomes of the interplay between personal characteristics and organizational factors in the workplace (3). Furthermore, quality of working life encompasses individual characteristics and organizational factors. At the opposite end of the spectrum, job involvement contrasts with job burnout. In other words, researchers consider the three dimensions of job involvement (vigor, dedication, and absorption) to be opposites of the three dimensions of burnout (emotional exhaustion, depersonalization, and lack of personal accomplishment) (43,44). Therefore, it can be hypothesized that quality of working life may influence job involvement through the mediation of burnout:

**H5:** Job burnout is negatively correlated with job involvement.

**H6:** Job burnout mediates the relationship between quality of working life and job commitment.

### 1.4. Job Identity and Its Relationship with Job involvement

There are numerous definitions of nurses’ job identity. According to Leddy & Pepper, job identity among nurses implies that they are able to perform the tasks involved in their roles while demonstrating good planning and ideals for nursing work (45). Öhlén describes the professional identity of nurses from both subjective and objective perspectives (46). The subjective aspect encompasses individual nurses’ feelings toward the nursing profession, whereas the objective aspect reflects an external view of nursing as a profession. In this study, job identity is defined as an individual’s perception of the social impact and significance of their work within a profession. Research on higher education staff suggests that job identity influences job commitment (47).

Additionally, a study on hotel employees indicates that job identity reduces employees’ intentions to leave their jobs (48). When employees believe that their work can provide them with personal value and meaning, they experience a higher sense of job identity, greater engagement, and increased job satisfaction. Conversely, when job identity is low, employees are more likely to leave their current positions if suitable opportunities arise (49,50).

**H7:** Job identity is positively correlated with job commitment.

This study aims to examine the impact of nurses’ quality of work life on their job involvement and the mediating pathways involved, specifically the intermediary roles of job identity and job burnout. Achieving this goal can contribute to the existing literature.

## 2. Methodology

This study is a cross-sectional research conducted in Khorramabad, Lorestan Province, Iran. The study population comprised nurses employed in hospitals in Khorramabad, with a total population size of 1,600. Using Cochran’s formula, a sample size of 310 nurses was determined, and participants were selected through simple random sampling. Nurses were included in the study if they met the following criteria: a minimum of two years of work experience and consent to participate in the research process. Exclusion criteria were a lack of willingness to continue participation for any reason. The instruments used in this study were the Work-Related Quality of Life Scale (WRQOLS), Maslach Burnout Questionnaire (1981), Professional Identity Scale developed by Hao et al. (2014), and the Utrecht Work Engagement Scale (UWES-9). Each of these tools is further described below.

### 2.1. Work-Related Quality of Life Scale (WRQOLS)

This questionnaire contains 34 items across 7 dimensions, including General Well-being (GWB), Work-home Interaction, Employee Engagement, Job and Professional Satisfaction, Workplace Stress, Working Conditions (WCS), Control at Work (CAW), rated on a 5-point Likert scale (1). Lin and colleagues validated the reliability and validity of this tool, with a Cronbach’s alpha coefficient of 0.88 (1). A validation study in the United Kingdom by Van Laar and colleagues confirmed that this scale has satisfactory reliability and validity for both the overall scale and its subscales (Cronbach’s alpha=0.86) (51).

### 2.2. Maslach Burnout Questionnaire (1981)

This questionnaire includes 22 questions designed to assess emotional exhaustion, depersonalization, and lack of personal accomplishment within a professional context. It is particularly used to track burnout phenomena in professional groups such as nurses, teachers, etc. This questionnaire was validated by Kochi (2015) (Cronbach’s alpha=0.91) (52). The dimensions of this questionnaire include emotional exhaustion, depersonalization, and personal accomplishment.

### 2.3. Professional Identity Questionnaire by Hao et al. (2014)

This questionnaire consists of 17 questions that assess 5 factors: professional self-image, career achievements, social comparison and self-reflection, professional autonomy, and social modeling. Responses are recorded on a 5-point Likert scale, ranging from strongly disagree (score=1) to strongly agree (score=5) (53). The validity and reliability of this tool have been confirmed, with a Cronbach’s alpha of 0.83 (53).

### 2.4. Utrecht Work Engagement Scale (UWES-9)

This questionnaire consists of 9 items, aiming to assess job involvement across multiple dimensions (vigor, dedication, absorption). The scoring is based on a 5-point Likert scale. In the study by Ghanbari et al., the face and content validity were assessed and confirmed by experts and faculty members. The Cronbach’s alpha was calculated to be 0.87 (54).

Structural Equation Modeling using Partial Least Squares (PLS-SEM) differs from covariance-based methods, as it lacks model fit indices based on chi-square tests to assess the alignment between the theoretical model and the collected data. This is due to the predictive nature of PLS. Therefore, the fit indices developed with this approach are related to evaluating the adequacy of the model in predicting dependent variables, such as redundancy and parsimony indices or the Goodness of Fit (GOF) index. These indices indicate how well the measurement model predicts its underlying constructs, and in the structural model, they show how well and with what quality the exogenous variables predict the endogenous variables. Thus, in this study, to estimate the relationships between variables, Structural Equation Modeling and Smart-PLS software were used.

## 3. Results

In this section, the model derived from the qualitative phase has been distributed among the nurses in the study using a questionnaire with a five-point Likert scale. This phase, which utilized structural equation modeling (SEM), was conducted to validate the model obtained in the qualitative phase. Therefore, in this study, the goodness-of-fit indices of the model were first reported, followed by the final results of the relationships between the variables. Figure 1 illustrates the model fit of the research.

**Figure 1:**
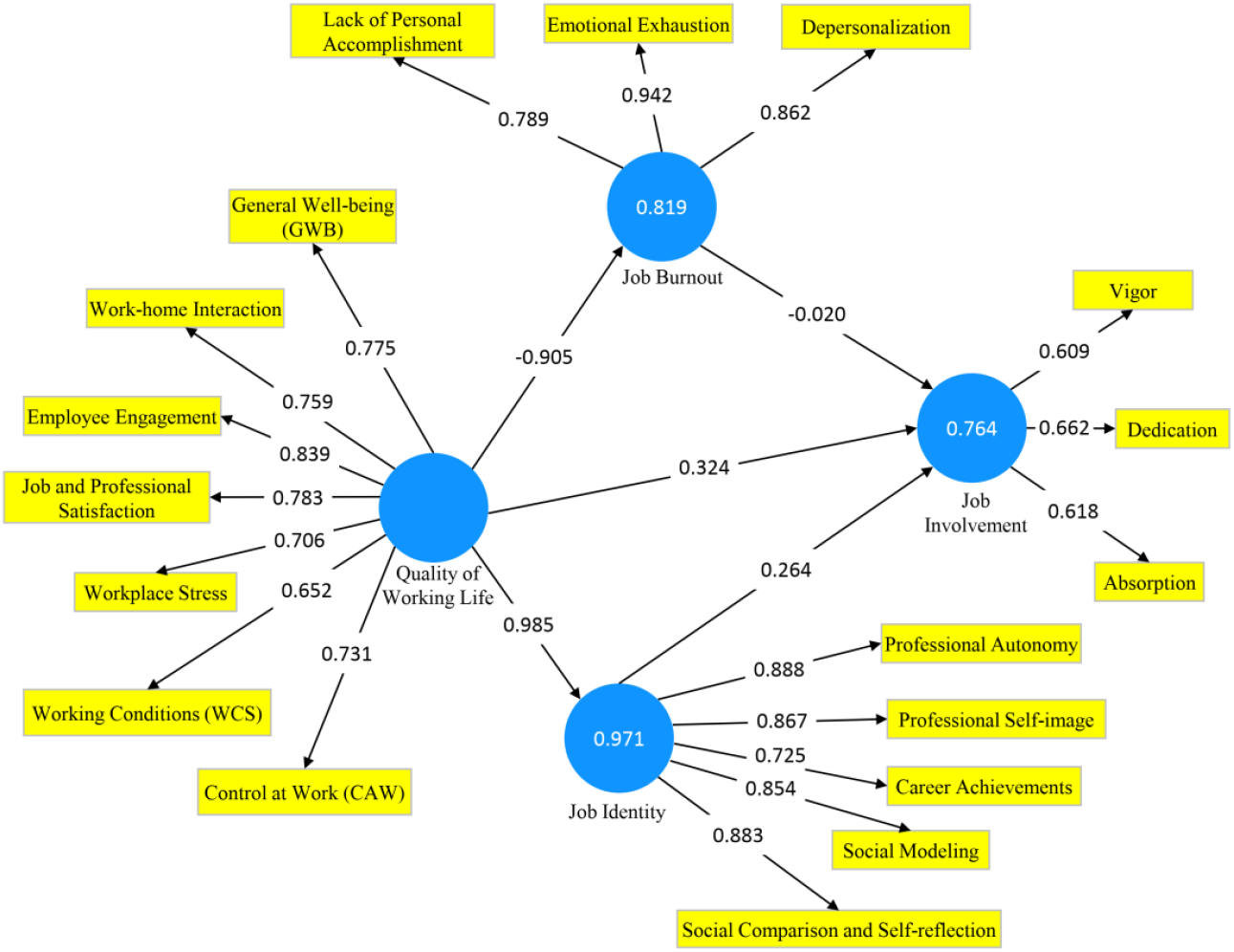
Model Fit (Path Coefficients)

### 3.1. Factor Loadings

In the case of a reflective measurement model, the model will be homogeneous if the absolute value of the factor loading for each observed variable corresponding to its latent variable is at least 0.60. Based on the results in Table 1, the factor loadings for all observed variables are greater than 0.60. Therefore, it can be concluded that the observed variables corresponding to the latent variables have been able to adequately explain their respective constructs.

**Table 1:** Factor Loadings of the Study Variables.

| Row | Latent Variable | Observed Variable | Factor Loading |
| --- | --- | --- | --- |
| 1 | Quality of Working Life | General Well-being (GWB) | 0.775 |
|  |  | Work-home Interaction | 0.759 |
|  |  | Employee Engagement | 0.839 |
|  |  | Job and Professional Satisfaction | 0.783 |
|  |  | Workplace Stress | 0.706 |
|  |  | Working Conditions (WCS) | 0.652 |
|  |  | Control at Work (CAW) | 0.731 |
| 2 | Job Identity | Professional Autonomy | 0.888 |
|  |  | Professional Self-image | 0.867 |
|  |  | Career Achievements | 0.725 |
|  |  | Social Modeling | 0.854 |
|  |  | Social Comparison and Self-reflection | 0.883 |
| 3 | Job Involvement | Vigor | 0.609 |
|  |  | Dedication | 0.662 |
|  |  | Absorption | 0.618 |
| 4 | Job Burnout | Depersonalization | 0.862 |
|  |  | Emotional Exhaustion | 0.942 |
|  |  | Lack of Personal Accomplishment | 0.789 |

### 3.2. Reliability and Validity Indices

Reliability indices include Cronbach’s alpha and composite reliability (p Deleuwin-Goldstein), with an acceptable threshold for these indices set at 0.7. According to the results in Table 2, Cronbach’s alpha and the composite reliability index (CR) for all latent variables exceed 0.7. The Cronbach’s alpha for the research variables is above 0.7, indicating that the reliability of the research variables is at a desirable level. Likewise, the composite reliability index for the research variables is also above 0.7, confirming a satisfactory level of reliability for these variables.

**Table 2:** Reliability and Validity of the Model.

| Variable | Cronbach's Alpha | CR | AVE |
| --- | --- | --- | --- |
| Job involvement | 0.750 | 0.764 | 0.697 |
| Job Burnout | 0.731 | 0.900 | 0.751 |
| Job Identity | 0.755 | 0.785 | 0.632 |
| Quality of Work Life | 0.790 | 0.849 | 0.653 |

To assess validity, the Average Variance Extracted (AVE) index was utilized. The Average Variance Extracted (AVE) criterion was proposed by Fornell & Larcker (1981) as a measure of the internal validity of reflective measurement models. This index represents the extent to which a construct correlates with its indicators. For this index, a minimum value of 0.5 is considered, meaning that the targeted latent variable explains at least 50% of the variance of its observed indicators. According to the results in Table 2, the AVE index for all latent variables is greater than 0.5, demonstrating that the validity of the research variables is at a desirable level.

### 3.3. Coefficient of Determination and Adjusted Coefficient of Determination

The primary criterion for assessing endogenous latent variables in a path model is the coefficient of determination. This indicator shows the percentage of variance in the endogenous variable that is explained by the exogenous variable. In structural path models, values of 0.19, 0.33, and 0.67 for endogenous latent variables are respectively described as weak, moderate, and substantial. However, if the endogenous latent variable is influenced by only a few exogenous variables, moderate values of the coefficient of determination are also acceptable. The adjusted coefficient of determination is similar to the coefficient of determination. The critical distinction between the coefficient of determination and the adjusted coefficient of determination is that the former assumes that each observed independent variable in the model explains the variance in the dependent variable. Thus, the percentage shown by the coefficient of determination assumes the influence of all independent variables on the dependent variable. In contrast, the percentage shown by the adjusted coefficient of determination results only from the actual influence of the independent variables in the model on the dependent variable, not from all independent variables. Another distinction is that the appropriateness of variables for the model may not be indicated by the coefficient of determination even with a high value, whereas one can rely on the estimated value of the adjusted coefficient of determination.

According to the results in Table 3, the coefficient of determination and adjusted coefficient of determination for the variable “job involvement” as the dependent variable are 0.764 and 0.758, respectively. Therefore, it can be said that the independent variables influencing job involvement explain 76.4% of the variance in this variable. The coefficient of determination and adjusted coefficient of determination for the variable “job burnout” as the dependent variable are 0.819 and 0.818, respectively. Therefore, it can be said that the independent variables influencing job burnout explain 81.9% of the variance in this variable. The coefficient of determination and adjusted coefficient of determination for the variable “job identity” as the dependent variable are both 0.971. Thus, it can be stated that the independent variables influencing job identity explain 97.1% of the variance in this variable.

**Table 3:**
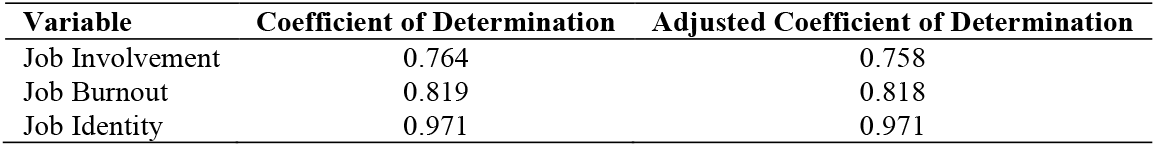
Coefficient of Determination and Adjusted Coefficient of Determination.

### 3.4. Effect Size F^2^

The F^**2**^ effect size index is utilized to determine the strength of the relationship between latent variables in the model, initially introduced by Cohen (1981). This metric enables the measurement of the effect size of an exogenous variable on an endogenous variable within structural equation modeling. Cohen suggested thresholds of 0.02, 0.15, and 0.35 to indicate small, medium, and large effect sizes, respectively. Based on the results presented in Table 4, the research variables exhibit strong effects on one another.

**Table 4:** F^2^ Effect Size Index.

| Variable | Job Involvement | Job Burnout | Job Identity | Quality of Work Life |
| --- | --- | --- | --- | --- |
| Job Involvement | - | - | - | - |
| Job Burnout | 0.262 | - | - | - |
| Job Identity | 0.720 | - | - | - |
| Quality of Work Life | 0.704 | 4.525 | 33.384 | - |

### 3.5. Cross-validated communality index (CV Com), cross-validated redundancy index (CV Red), and the Goodness-of-Fit (GOF)

Another criterion for evaluating the measurement model is the test for assessing its quality. The quality of the measurement model is assessed through the cross-validated communality index (CV Com), cross-validated redundancy index (CV Red), and the Goodness-of-Fit (GOF) index. If the communalities and redundancy indices are positive, it indicates that the measurement model possesses the necessary quality. Additionally, the GOF index, which is the square root of the product of the average communality values and the average coefficient of determination, ranges between zero and one. Wetzels et al. (2009) defined values of 0.01, 0.25, and 0.36 as weak, moderate, and strong GOF thresholds, respectively. According to the results in Table 5, the communalities and redundancy indices for the latent variables are positive. Moreover, the GOF index, with a value of 0.471, indicates that the fitted model demonstrates satisfactory quality.

**Table 5:** Quality Indices of the Measurement Model.

| Variable | CV Com | CV Red | GOF |
| --- | --- | --- | --- |
| Job Involvement | 0.103 | 0.130 | 0.471 |
| Job Burnout | 0.465 | 0.580 |  |
| Job Identity | 0.192 | 0.394 |  |
| Quality of Work Life | 0.285 | - |  |

### 3.6. Direct Effects Estimation

According to the results in Table 6, job burnout has a direct, negative, and significant impact on job involvement, with an estimated coefficient of -0.020, which is significant at the 99% confidence level. Job identity exerts a direct, positive, and significant impact on job involvement, with an estimated coefficient of 0.264, significant at the 99% level. Quality of work life has a direct, positive, and significant effect on job involvement, with an estimated coefficient of 0.324, significant at the 99% confidence level.

**Table 6:** t-test of Direct Effects.

| Variable | Sample Value | Sample Mean | Standard Deviation | T-Statistic | Significance Level |
| --- | --- | --- | --- | --- | --- |
| Job Burnout → Job involvement | -0.020 | -0.047 | 0.003 | 6.103 | 0.000 |
| Job Identity → Job involvement | 0.264 | 0.351 | 0.030 | 8.650 | 0.000 |
| Quality of Work Life → Job involvement | 0.324 | 0.211 | 0.043 | 7.603 | 0.000 |
| Quality of Work Life → Job Burnout | -0.905 | -0.907 | 0.021 | 42.649 | 0.000 |
| Quality of Work Life → Job Identity | 0.985 | 0.985 | 0.004 | 278.770 | 0.000 |

Furthermore, quality of work life has a direct, negative, and significant effect on job burnout, with an estimated coefficient of - 0.905, significant at the 99% level. Quality of work life also has a direct, positive, and significant impact on job identity, with an estimated coefficient of 0.985, significant at the 99% confidence level.

### 3.7. Indirect Effects Estimation

Based on the results in Table 7, quality of work life, through job burnout, has a positive and significant effect on job involvement, with an estimated coefficient of 0.018, significant at the 99% confidence level. This suggests that job burnout serves as a mediating variable between quality of work life and job involvement. Quality of work life also has a positive and significant impact on job involvement through job identity, with an estimated coefficient of 0.261, significant at the 99% level, indicating that job identity acts as a mediator between quality of work life and job involvement.

**Table 7:**
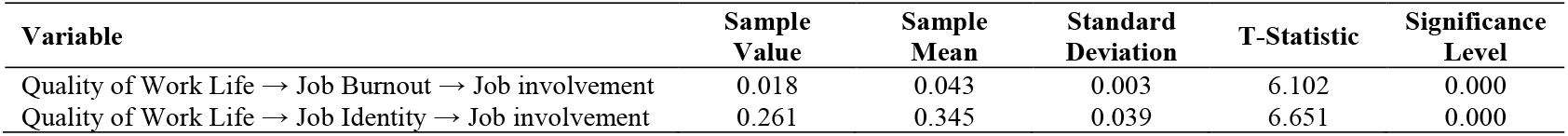
t-test of Indirect Effects.

### 3.8. Fitting the Total Effects

Based on the results in Table 8, job burnout has a significant negative impact on job involvement. The estimated coefficient is - 0.020, which is significant at the 99% level. Job identity has a significant positive effect on job involvement, with an estimated coefficient of 0.264, also significant at the 99% level. Quality of work life positively and significantly influences job involvement, with an estimated coefficient of 0.602, significant at the 99% level. Additionally, quality of work life has a negative and significant effect on job burnout, with an estimated coefficient of -0.905, significant at the 99% level. Quality of work life has a positive and significant effect on job identity, with an estimated coefficient of 0.985, also significant at the 99% level.

**Table 8:** Total Effects t-test.

| Variable | Sample Value | Sample Mean | Standard Deviation | T-Statistic | Significance Level |
| --- | --- | --- | --- | --- | --- |
| Job Burnout → Job Involvement | -0.020 | -0.047 | 0.003 | 6.103 | 0.000 |
| Job Identity → Job Involvement | 0.264 | 0.351 | 0.030 | 8.650 | 0.000 |
| Quality of Work Life → Job Involvement | 0.602 | 0.600 | 0.066 | 9.167 | 0.000 |
| Quality of Work Life → Job Burnout | -0.905 | -0.907 | 0.021 | 42.249 | 0.000 |
| Quality of Work Life → Job Identity | 0.985 | 0.985 | 0.004 | 278.770 | 0.000 |

### 3.9. Pearson Correlation Analysis

According to the results in Table 9, there is a significant and negative correlation between job involvement, job identity, and quality of work life with job burnout. The correlation coefficients are -0.910, -0.878, and -0.900, respectively.

**Table 9:** Pearson Correlation Test.

| Variable | Job Involvement | Job Burnout | Job Identity | Quality of Work Life |
| --- | --- | --- | --- | --- |
| Job Involvement | 1 | -0.910 | 0.953 | 0.968 |
| Job Burnout | -0.910 | 1 | -0.878 | -0.900 |
| Job Identity | 0.953 | -0.878 | 1 | 0.992 |
| Quality of Work Life | 0.968 | -0.900 | 0.992 | 1 |

## 4. Discussion

This study utilized a cross-sectional survey of 310 nurses to examine the relationship between quality of work life (QWL) and job involvement in nursing. It also tested the mediating roles of burnout and job identity using a structural equation model designed to address gaps in the existing literature. One critical aspect of QWL is work-life balance. Thus far, various approaches have been introduced to foster this balance. For instance, Denton explored the concept of “flexible working hours” (55), emphasizing that systematic implementation is necessary for improving job efficiency and employee morale. Similarly, numerous studies highlight that the return on investment in work-life projects can be significantly positive, with the primary outcome being the retention of human capital within organizations (56). Some research suggests that as nurses age, job involvement tends to decline (57). Patience et al. proposed a prospective model addressing nurses’ perspectives on job demands and resources for engagement, based on the Job Demands-Resources Theory. They subsequently recommended strategies to enhance nurses’ work engagement from the perspective of job demands and resources (58).

This study not only creatively explores the relationship between QWL and job involvement but also reveals the pathway of influence between them. The findings indicate that job identity and burnout mediate the relationship between QWL and job commitment. Results suggest that when nursing managers aim to enhance job involvement among new nurses, they should consider not only QWL but also the aspects of job identity and burnout. Social Identity Theory posits that identity can impact individual cognition, emotions, and behavior (59). Job identity among nurses is an integrative process of cognition, emotion, and behavior, ultimately influencing individual nursing behaviors. Nurses with high job identity tend to experience positive feelings toward nursing, exhibit greater pride and efficiency in their work, achieve a heightened sense of accomplishment and well-being, and are better able to engage in their work with increased enthusiasm.

Some researchers propose that the most effective interventions are those combining specific actions at both organizational and individual levels (60). Based on the Job Demands-Resources Theory, Bakker et al. suggest two types of interventions to prevent burnout and promote job involvement: organizational and individual interventions (44). Organizational-level interventions encompass optimizing job demands, increasing job resources, and fostering personal resources. Individual-level intervention implies that the organization can address specific needs and challenges that employees may face, such as 1) job crafting training (61), 2) strengths use training, in which employees learn to set personal goals and apply their strengths in new ways at work (62), and 3) recovery training (63). Recovery practices might involve relaxation techniques or mindfulness exercises. Consequently, training programs can be established to enable new nurses to better cope with the challenges and pressures of their new roles. Furthermore, the results of this study explain how burnout may be managed by providing essential QWL features, such as fair and equitable wages, safe and healthy working conditions, social integration within the workplace, and work-life social connectivity. Some researchers suggest that enhancing QWL features could increase psychological empowerment and reduce nurses’ burnout (25,64). Additionally, professional training and improving nurses’ communication skills help them manage burnout more effectively (65).

## 5. Conclusion

The findings from this study affirm the robustness of the proposed model through structural equation modeling (SEM), which indicated a satisfactory model fit and reliable relationships among variables. The high factor loadings, exceeding 0.60 across all observed variables, demonstrate the model’s capacity to capture each construct effectively. Additionally, reliability and validity indices such as Cronbach’s alpha, composite reliability, and Average Variance Extracted (AVE) meet recommended thresholds, underscoring the consistency and validity of the measurements. Moreover, the results highlight significant direct and indirect effects among the variables, particularly showing how quality of work life (QWL) positively impacts job identity and involvement, while inversely affecting job burnout. The cross-validated communality and redundancy indices, alongside a strong Goodness-of-Fit (GOF) index of 0.471, further confirm the model’s adequacy. The presence of significant mediating roles of job identity and burnout in the relationships between QWL and job involvement illustrates complex interactions, revealing that enhancing QWL could lead to improved job involvement and reduced burnout. This comprehensive model provides critical insights into factors that influence nurses’ professional engagement and well-being, potentially guiding future interventions.

## Data Availability

All data produced in the present study are available upon reasonable request to the authors

## Acknowledgements

We would like to express our sincere gratitude to all those who supported and collaborated with us in conducting this research. We are deeply thankful to our esteemed professors, research team members, and friends, whose valuable guidance contributed to the quality of this study. We are also grateful for the financial support and resources provided by Lorestan University of Medical Sciences, without which this research would not have been possible.

## Author Contribution

All authors contributed actively to the preparation of this manuscript. **E.N**. was responsible for the study design and data collection. **A.R**. participated in data analysis and drafting the initial manuscript. **T.S**. reviewed and edited the manuscript and provided overall supervision. All authors have read and approved the final version of the manuscript.

## Ethical Considerations

This research was conducted in full compliance with ethical research principles and was approved by the Institutional Review Board (IRB) at the Lorestan University of Medical Sciences (Ethics code: IR.LUMS.REC.1402.266). Data were collected using a questionnaire designed to gather the opinions of nurses. Informed consent was obtained from all participants, who voluntarily agreed to participate after being fully informed about the study’s purpose and the use of their information. All collected data were kept confidential and used solely for research purposes. Participants had the right to withdraw from the study at any time without any negative consequences.

